# The Effectiveness of a Whole System Approach to Improve Physical Activity of Children Aged 5 to 11 Years Living in Multi-ethnic and Socio-economically Deprived Communities: A Controlled Before and After Trial

**DOI:** 10.1101/2025.03.20.25324322

**Authors:** SE Barber, DD Bingham, NP Dawkins, Z Helme, J Hall, A Seims, G Santorelli, J Wright, RRC McEachan, J Burkhardt, A Daly-Smith

## Abstract

**Introduction:** Whole system approaches to public health challenges such as low physical activity levels have the potential to create sustained behaviour change at a population level and tackle health inequalities. However, there is currently little evidence of the nature or effectiveness of adopting whole system approaches. This study evaluated whether a whole system physical activity intervention (JU:MP), was effective at improving physical activity in five- to eleven-year-olds.

**Methods:** A before and after controlled study with two-arms (JU:MP intervention and control), was conducted in Bradford, UK with data collected at baseline and 24-months follow-up. Habitual physical activity was measured via accelerometry. The primary outcome was difference in moderate-to-vigorous intensity physical activity (MVPA) between groups at 24-months. Secondary outcomes included: sedentary time (ST), counts per minute (CPM), BMI z-score, waist circumference, social, emotional and behavioural health, and quality-of-life. An exploratory analysis compared intervention effects between sub-groups.

**Results:** 1,453 children were recruited. 330 children with valid wear-time at baseline and 24- months (JU:MP group n=175, control group n =155) were included in the final analysis of physical activity outcomes. The JU:MP group improved levels of MVPA (+4.99 minutes/day, (CI = 1.01, 8.96), standardised mean difference (SMD) = 0.29), ST ( -8.69 minutes/day, CI = -16.76, -0.61), SMD = -0.20) and CPM (+32.72, CI = 5.93, 59.53, SMD = 0.28) compared to controls. There were minor differences between groups in all secondary outcomes, favouring the JU:MP group. Exploratory sub-group analysis revealed that MVPA improved for boys (+7.34 minutes/days, CI = 0.70, 13.99, SMD = 0.36) and South Asian heritage children (+7.20 minutes/day, CI = 1.67, 12.72, SMD = 0.52) in the JU:MP group compared to the control group. Conclusion: whole system approaches hold considerable promise for addressing children’s levels of physical activity at scale, whilst also tackling inequalities.

**Key messages:** *What is already known on this topic:* The physical activity levels of children are influenced by complex political, environmental and social systems. The World Health Organisation and the International Society of Physical Activity and Health both advocate for whole system change to support population level improvements in physical activity.

*What the study adds:* This study provides evidence that improving population levels of physical activity in children can be achieved by taking a whole system approach.

*How this study might affect research practice or policy:* This study can give confidence to policy makers and practitioners who are considering or continuing to take a whole system approach to improve physical activity for populations at greatest need.

## Introduction

Improving children’s levels of physical activity is a high priority for public health. Most (∼70%) of the children in the UK do not meet the Chief Medical Officers guidelines of an average of 60 minutes moderate-to-vigorous intensity physical activity (MVPA) a day (1). There are inequalities in activity levels, with children from black and Asian ethnic minority backgrounds and those from the least affluent families less active than other ethnicity and affluence groups (2). Girls are also less active than boys across all ages in childhood (2). Undertaking sufficient daily physical activity and reducing sedentary time is beneficial for children’s physical, social and emotional health, their growth and development and prevents the early onset of diseases(3). Physical activity behaviours typically decline across childhood and adolescence from age 6-7 through to age 14-15 and they track from childhood, through adolescence and into adulthood (4). Establishing adequate physical activity behaviours and mitigating this decline can have life-long benefits for health and wellbeing. In addition to the health-related consequences of physical inactivity, it is estimated the economic cost of inactivity in the UK is £7.4 billion each year (5), which emphasises the urgent need to address this issue.

The physical activity and movement behaviours of populations are influenced by complex political, environmental and social systems rather than just an individual’s ‘intention’ to be active. Whole system approaches that intervene in these complex systems to improve population level physical activity are advocated for by the World Health Organisation and the International Society of Physical Activity and Health (6,7). Authentic whole system approaches draw on complexity science and complex adaptive systems (8). They should contain, 1) heterogeneous interacting elements, 2) an emergent effect that is different from the effects of the individual elements, and 3) persisting effects over time that adapt to changing circumstances(9). Multi-faceted interacting interventions which work at various ‘levels’ of a whole system (e.g. individuals, communities, organisations, environment and policy and strategy) (9,10), have the potential to lead to sustained behaviour change at a population level (9,10) and may help to reduce health inequalities. Despite system approaches to physical activity being increasingly advocated for, evidence of their operationalisation and effectiveness is sparse.

A scoping review by Nau et al. identified seven studies which focused on system approaches for adult physical activity (11). There were no whole system approaches to improve children’s physical activity per se, but four designed for childhood obesity prevention, which included some elements targeting physical activity behaviours. Nau et al. concluded that this field of research is at an early stage of development, with most literature reporting descriptives of approaches and providing no further complex analyses (11). A further systematic review by Koorts et al. found eight interventions with high or moderate use of systems approaches to drive impact at scale to tackle non-communicable diseases, which included a physical activity component (12). Only one targeted children (13); it took a whole-of-school approach (e.g. a school system approach) rather than targeting multiple settings across a whole ‘local’ system (e.g. children, families, the wider community, schools, community organisations, faith settings, environments including green spaces and active travel infrastructure, local government policy and strategy) and was deemed as having moderate use of a systems approach.

Since 2018, Sport England (a non-departmental public body of the UK government, which funds and supports people and communities across England to be active) has funded twelve localities across England to pilot whole system, place-based approaches to reduce physical inactivity and health inequalities (14). One of these localities was Bradford, where the approach, called JU:MP, aimed to improve physical activity in children aged five to fourteen years living in some of the most deprived areas of England, with a focus on tackling physical activity inequalities for girls and for ethnic minority groups (15).

The primary aim of this study was to explore whether JU:MP; a whole systems community-based physical activity approach delivered at scale, was effective at improving MVPA among children aged five to eleven years old. Secondary aims were to determine the effectiveness of the intervention on impacting broader measures of physical behaviours, adiposity, social emotional and behavioural health, and quality of life.

## Materials and Methods

### The JU:MP intervention

The JU:MP intervention took a theory and evidence based approach (drawing on complex adaptive systems thinking, the socio-ecological model, COM-B behaviour change theory, findings from the DEDIPAC reviews (16–19), and local evidence (20) coupled with community co-design and co-production. It worked across and linked five elements of the ‘local system’ (policy and strategy, environment, communities, organisations and children and families).

Sixteen work streams that sat within at least one element of the ‘local system’ (and often multiple) were developed and implemented in eight geographically defined neighbourhoods across the north of Bradford city reaching 30,000 children. The intervention aimed to increase capability, opportunity and motivations for physical activity in the targeted communities. This included (but was not limited to) creating safer more appealing play spaces, increasing opportunities for active transport, creating a culture for physical activity in schools, faith settings, and community organisations, and educating and motivating families and community members through physical activity social marketing campaigns. The different intervention elements interacted with each other via various mechanisms including staff whose role it was to connect across the system and local neighbourhood action groups whereby partners from across the system regularly came together to design and deliver a local action plan. An iterative learning approach was taken to understand what worked (and did not work) for who, how and why; this supported the continuous development of JU:MP and allowed it to adapt to the changing social, political and economic context. In each neighbourhood JU:MP had an intensive 24 month delivery phase followed by a 6 month embed and sustain phase in which financial support for activities tailed off and activities were supported to self-sustain. A more detailed description of the JU:MP whole system approach has been published elsewhere (21) as can resources relating to the intervention (22). The JU:MP intervention was piloted in three neighbourhoods between 2019-2021, after this the intervention was delivered in a further five neighbourhoods in 2021-2024. Three of the 2021-2024 neighbourhoods were used to conduct the effectiveness study.

### Study Design

The effectiveness of the JU:MP intervention was assessed using a two-arm (JU:MP intervention and control), non-blinded, non-equivalent group before and after study design; outlined in our published protocol (23)(ISRCTN14332797). This design included three waves of data collection (baseline [before JU:MP intervention], 24 months [at the end of the intensive 24 month delivery phase], and 36 months [12 months after the intensive activities were completed]), with this study focusing on data from the baseline and 24 month time-points. The study and all processes were given ethical approval by the University of Bradford Research Ethics Committee (Humanities, Social, and Health Sciences Research Ethics Panel; reference: (E891, July 2021).

### Patient and Public Involvement and Engagement (PPIE)

During conception of the study design and methodology we consulted with the Born in Bradford Parental Involvement group and the Born in Bradford Young Ambassadors group (children were aged 9-12 years old) to explore measurement tools and protocols that would be feasible and acceptable for children and parents. We attended a Bradford Schools Linking Network meeting as part of the Centre for Applied Education Research (24) and discussed with teachers and school senior leadership staff how the study could be delivered in schools with minimal disruption to the school day. After the baseline data collection we held some small group discussions with children who had taken part in the data collection to understand what would improve their experience of participation for the 24 month follow-up, as a result of this we employed privacy screens to measure waist circumference during follow-up measures. To engage with study participants throughout the study we created and shared child-friendly animations of the different aspects of the study (e.g. what happened during data collection, what happened to their data after it had been collected and a summary of the findings of the study).

### Sampling and recruitment of school and participants

The JU:MP intervention group was recruited from three neighbourhoods selected to ensure that children with White British ethnicity and children from ethnic minority groups were represented in the trial. Control neighbourhoods were selected from within Yorkshire and matched to the JU:MP intervention neighbourhoods (as according to the protocol) to ensure socio-demographic comparability. The matching was based on school-level Index of Multiple Deprivation (IMD) (25)(maximum 1 decile difference), free school meal eligibility (maximum 10% difference), and predominant ethnic group composition (maximum 15% difference). Within the selected areas, government-funded primary schools were invited to participate in the study, with incentives offered for participation. All children in Years one, two, and three (ages five to eight years) at recruited schools were invited to participate. Parents/carers provided informed consent for their child to participate before data collection. On the day of data collection, each child provided verbal assent, with appropriate support provided for those children with special educational needs/ disabilities. All children received a gift “goodie bag” at each data collection time-point.

At baseline, schools provided demographic data including child date of birth, biological sex, ethnicity, home postcode (for IMD calculation), child’s disability or special educational needs, and receipt of government-funded free school meals for each participant.

### Outcome Measures

At each time-point the following measures were collected for children unless otherwise stated: accelerometer assessed physical activity, anthropometric data, social emotional and behavioural health (teacher rated), and questionnaire assessed physical activity, sedentary behaviour, sleep, and quality of life (parent/carer rated).

#### Accelerometer assessed physical activity

Physical activity and sedentary time were measured using waist-worn ActiGraph accelerometers placed on the participant’s right hip, MVPA was the primary outcome variable. Participants were asked to wear the accelerometer for seven consecutive days (24-hours a day). The ActiGraph data was processed using ActiLife v6 (ActiGraph) and downloaded in 60- second epoch files. Sleep was identified using the validated sleep period–detection algorithms (26,27), and removed from the physical activity analysis. Physical activity intensities were classified using the Evenson cut points (28). To ensure consistency and increase validity, data was reprocessed using software Actilife v6, to generate 15-second epoch data to align with Evenson’s protocol (29). From this, data were categorised to establish the following outcome variables: sedentary time (ST), light physical activity (LPA), moderate-to-vigorous physical activity (MVPA), total physical activity (TPA), and average daily counts per minute (CPM). Nonwear time was defined as 20 minutes of consecutive zeros and was removed from the data. The wear-time criteria used was 600 minutes of wear to establish a valid day. Participants required three days of wear time, including at least one weekend day, at both baseline and follow-up, to be included in the analysis. This wear-time criterion was calculated using published local Bradford data and has an estimated intraclass correlation of 0.75 (30,31).

#### Anthropometric data

Anthropometric measurements were collected during data collection visits at each time point, with each measure repeated three times. Participants were requested to remove shoes and jumpers for each anthropometric measurement. Height (cm) and mass (kg) were measured, using a portable stadiometer (Seca 213, Hamburg, Germany and digital scales (Seca 875, Hamburg, Germany), to the nearest 0.1 cm and 0.1 kg respectively. BMI was calculated and converted to a BMI percentile and z-score based on UK reference data (32). Waist circumference (cm) was measured to the nearest 0.1cm at the narrowest point between the bottom rib and the iliac crest, in the midaxillary plane, using an ergonomic circumference tape (Seca 201 Hamburg, Germany).

#### Children’s social emotional and behavioural health (teacher rated)

Social, emotional and behavioural health was assessed by class teachers using the strengths and difficulties questionnaire (SDQ)(33). The SDQ contains 25 items, grouped into 5 subscales: “prosocial behaviour”, “emotional problems”, “behavioural problems”, “peer problems”, and “hyperactivity/inattention”.

#### Questionnaire assessed physical activity, sedentary behaviour, sleep, and quality of life (parent/carer rated)

Parents/carers completed a questionnaire to assess their child’s physical activity, sedentary behaviour, sleep and quality of life. The questionnaire consisted of six sections which covered: 1) personal information; 2) The Youth Activity Profile; 3) sleep duration; 4) parent-reported physical activity in specific settings; 5) parental practices; and 6) quality of life. Section one, personal information included: school class, teacher’s name, age and the relationship to the person completing the questionnaire. Section two included the Youth Active Profile, a previously published physical activity questionnaire which entails reporting the frequency and duration of physical activities engaged in through segments of a usual day (34). The Youth Activity Profile was also used to assess sedentary behaviours over the previous seven days. Section three collected the time the children went to bed and woke up on weekdays and weekends. These data were used to calculate average sleep time (35). Section four assessed the children’s parent-reported physical behaviours. This section included questions on: the frequency and locations where children were physically active for at least 10 minutes over the last 7 days; and how frequently children were physically active in parks and green spaces. In addition, if the child was of Muslim faith, parents were asked to complete a subsection including questions on the frequency of attendance at a Mosque or Madrasa; the duration of stay; and whether there was any active travel to or from the Mosque or Madrasa. Section five included questions focussed on physical behaviours and neighbourhood walkability. These questions were taken from previously validated questionnaires (36,37). Section six included questions on the child’s quality of life, taken from the parent version of the EQ-5D-Y and PedsQL (38–40).

### Statistical Analysis

Power calculations are detailed in the published protocol (23). Briefly, the calculation accounted for 6 clusters (3 JU:MP intervention and 3 control neighbourhoods), with a 5% two-sided alpha, and based on pilot data, an assumed control mean of 53.7 minutes of MVPA per day, a standard deviation of 19.7, and an intracluster correlation (ICC) rounded to 0.01 was used. To detect a 10-minute difference in the primary outcome of average daily MVPA, with 80% power at 24-month follow-up, accounting for 30% baseline accelerometer noncompliance and 50% follow-up data attrition; a minimum sample of 1,200 children (600 per condition group) was recommended, leaving a minimum final sample of 350 children (175 per condition group). Mixed effects regression models assessed JU:MP intervention effects on physical activity and sedentary outcomes, accounting for individual and group-level factors. The primary outcome was the difference between control and JUMP groups average daily MVPA at 24 months follow-up, while secondary outcomes included differences between groups average daily ST, LPA, TPA, and CPM. Random intercepts were included for clustering effects at the school and neighbourhoods levels (41). Given the relatively small final sample of participants with both valid baseline and follow-up data (n=330) and the natural skewness of accelerometer data (42,43); bootstrapping with 10,000 replications was used to enhance robustness, providing robust standard errors and confidence intervals (44). Bootstrapping has been used in physical activity research to improve estimate stability in cases of skewed data or limited sample sizes, as recommended in general methodological literature (44) and applied in previous physical activity data analyses (45–48). Standardised mean differences (SMD) measured effect sizes and were interpreted as small (0.2 to 0.5), medium (0.5 to 0.8), or large (≥0.8)(49). Adjusted means for the JU:MP intervention and control groups were calculated using post-hoc predictive margins. All models were adjusted for covariates, including sex, ethnicity, baseline values, wear time, BMI z-score, age, receipt of free school meals, school, and neighbourhood. Analyses followed the intention-to-treat principle, incorporating all participants with valid baseline and follow-up data to maintain group assignment and minimize potential biases (50). All analyses were conducted in STATA v.17, and statistical significance was set at p < 0.05.

## Results

### Recruitment and Control Identification

Figure 1 outlines the flow chart of schools and participants. Between June and December 2021, 95 primary schools across the Yorkshire region in England were approached; 17 out of the 19 intervention schools approached (89%), and 20 out of the 79 control schools approached (26.3%) agreed to participate. Control neighbourhoods were closely matched on the number of eligible pupils and the ethnicity of pupils (less than 5% difference between pupils who were classed as White British and South Asian). However, control neighbourhoods had a lower proportion of children eligible for free school meals (30.7% v 26.6%), and a higher median school IMD decile score (1 v 2) (refer to table/supplementary file for school and neighbourhood characteristic).

Of 4,408 eligible children (school year group 1 to 3), 1,453 parents/carers provided consent (34.2% JU:MP intervention, 31.7% control). A total of 316 children (21.7%) dropped out of the study at 24 months follow-up (T1), reasons being, school withdrawal (1 JU:MP school, 2 control schools), leading to n=99 (6.8%), dropping out. A further 217 (14.9%) children dropped out due to absence/illness during data collection. For accelerometry, 800 (55.1%) children met the wear-time criteria at baseline, 505 (34.8%) at 24 months, leaving a final sample of 330 (22.7%) which met the wear-time criteria at both time points and were included in the primary outcome analysis and analysis of other physical activity variables. For other secondary outcomes a total of 1,103 (75.9%) children had anthropometric data, 702 (48.3%) parent questionnaire data and 716 (49.3%) teacher questionnaires at both baseline and follow-up (Figure 1).

#### Demographics

Baseline demographic characteristics are presented in Table 1. Intervention (n=766) and control (n=687) groups showed no differences in sex but differed in ethnicity and socio-economic indicators at baseline (n=1,453). The JU:MP intervention group had fewer white British participants (38.9% vs. 44.4%) and higher free school meal eligibility (30.6% vs. 25.9%). Deprivation was greater in the JU:MP group, with 72.2% of children in the most deprived quintile versus 58.1% in controls. Physical activity outcomes showed minor differences, with the JU:MP group exhibiting higher light physical activity (LPA) and total physical activity. Sedentary time and MVPA did not differ between groups. Descriptive analysis compared the final analysis sample (n=330) with participants excluded from the main analysis due to not meeting accelerometer wear time criteria (n=1,123), see Table 2. Both groups had a similar male-to-female ratio and comparable ethnic composition. However, socioeconomic differences were observed; fewer participants in the final sample were eligible for free school meals, and fewer were from the most deprived quintile. More final sample participants resided in Neighbourhood 3, age and BMI Z-scores were similar across groups.

**Table 1.** Demographic descriptive statistics of recruited sample and baseline accelerometer data.

| Total Sample: n=1453 |  | Intervention |  | Control |  | P-Value |
| --- | --- | --- | --- | --- | --- | --- |
|  |  | n=766 (52.7%) |  | n=687 (47.3%) |  |  |
|  |  | N / Median | % / IQR | N / Median | % / IQR |  |
| Sex |  |  |  |  |  |  |
| Female |  | 411 | 53.66 | 357 | 51.97 | 0.572 |
| Male |  | 355 | 46.34 | 330 | 48.03 |  |
| Ethnicity |  |  |  |  |  |  |
| White British |  | 298 | 38.9 | 305 | 44.4 | 0.04 |
| South Asian |  | 342 | 44.65 | 295 | 42.94 |  |
| Other |  | 126 | 16.45 | 87 | 12.66 |  |
| Special Educational Needs or disability |  |  |  |  |  |  |
| Yes |  | 78 | 10.18 | 73 | 10.63 | 0.782 |
| No |  | 688 | 89.82 | 614 | 89.37 |  |
| Health condition |  |  |  |  |  |  |
| Yes |  | 128 | 16.71 | 570 | 17.03 | 0.871 |
| No |  | 638 | 83.29 | 117 | 82.97 |  |
| Parent/carer Consent |  |  |  |  |  |  |
| Mother |  | 598 | 78.07 | 549 | 80.03 | 0.162 |
| Father |  | 151 | 19.71 | 117 | 17.06 |  |
| Other |  | 17 | 2.22 | 21 | 2.91 |  |
| Free School Dinners |  |  |  |  |  |  |
| Yes |  | 234 | 30.63 | 178 | 25.91 | 0.047 |
| No |  | 530 | 69.37 | 509 | 74.09 |  |
| Index of Multiple Deprivation (Quintile) |  |  |  |  |  |  |
| 1 |  | 553 | 72.19 | 399 | 58.08 | 0.000 |
| 2 |  | 171 | 22.32 | 222 | 32.31 |  |
| 3 |  | 20 | 2.61 | 29 | 4.22 |  |
| 4 |  | 16 | 2.09 | 31 | 4.51 |  |
| 5 |  | 6 | 0.78 | 6 | 0.87 |  |
| Neighbourhoods |  |  |  |  |  |  |
| 1 |  | 236 | 30.81 | 208 | 30.28 | 0.877 |
| 2 |  | 282 | 36.81 | 262 | 38.14 |  |
| 3 |  | 248 | 32.38 | 217 | 31.59 |  |
| Age years |  | 6.78 | 6.07, 7.46 | 7.05 | 6.27, 7.73 | 0.000 |
| Body Mass Index Z-score |  | 0.382 | -0.442, 0.382 | 0.229 | -0.549, 1.202 | 0.0414 |
| MVPA average daily minutes* |  | 66.37 | 23.25 | 66.68 | 22.06 | 0.8434 |
| ST average daily minutes* |  | 387.21 | 58.98 | 392.98 | 46.15 | 0.1262 |
| LPA average daily minutes* |  | 327.63 | 53.59 | 315.18 | 43.59 | 0.0004 |
| Total PA average daily minutes* |  | 393.99 | 64.44 | 381.86 | 53.95 | 0.0042 |
| Wear time average daily minutes* |  | 781.20 | 50.83 | 774.85 | 49.33 | 0.0737 |
| Counts Per minute (CPM)* |  | 646.58 | 161.90 | 645.42 | 143.01 | 0.9149 |

\*Data for participants with valid data (800 participants at baseline)

**Table 2.** Demographic descriptive statistics of final analysis sample.

| Table 2 - Demographic descriptive statistics of final analysis sample |  |  |  |  |  |  |
| --- | --- | --- | --- | --- | --- | --- |
|  |  | Sample not included in main analysis<br>n=1123 |  | Main final Analysis Sample<br>n=330 |  | P-Value |
|  |  | N / Median | % / IQR | N / Median | % / IQR |  |
| Sex |  |  |  |  |  |  |
| Female |  | 603 | 53.7 | 165 | 50 | 0.237 |
| Male |  | 520 | 46.3 | 165 | 50 |  |
| Ethnicity |  |  |  |  |  |  |
| White British |  | 466 | 41.5 | 137 | 41.52 | 0.681 |
| South Asian |  | 497 | 44.26 | 140 | 42.42 |  |
| Other |  | 160 | 14.25 | 53 | 16.06 |  |
| Special Educational Needs or disability |  |  |  |  |  |  |
| Yes |  | 126 | 11.22 | 25 | 7.58 | 0.057 |
| No |  | 997 | 88.78 | 305 | 92.42 |  |
| Health condition |  |  |  |  |  |  |
| Yes |  | 186 | 16.56 | 59 | 17.88 | 0.575 |
| No |  | 937 | 83.44 | 271 | 82.12 |  |
| Parent/carer Consent |  |  |  |  |  |  |
| Mother |  | 872 | 77.72 | 275 | 83.33 | 0.377 |
| Father |  | 216 | 19.25 | 52 | 15.76 |  |
| Other |  | 35 | 3.03 | 3 | 0.91 |  |
| Free School Dinners |  |  |  |  |  |  |
| No |  | 779 | 69.49 | 260 | 78.79 | 0.001 |
| Yes |  | 342 | 30.51 | 70 | 21.21 |  |
| Index of Multiple Deprivation (Quintile) |  |  |  |  |  |  |
| 1 |  | 759 | 67.59 | 193 | 58.48 | 0.002 |
| 2 |  | 276 | 24.58 | 117 | 35.45 |  |
| 3 |  | 42 | 3.74 | 7 | 2.12 |  |
| 4 |  | 37 | 3.29 | 10 | 3.03 |  |
| 5 |  | 9 | 0.8 | 3 | 0.91 |  |
| Neighbourhoods |  |  |  |  |  |  |
| 1 |  | 358 | 31.88 | 86 | 26.06 | 0.001 |
| 2 |  | 433 | 38.56 | 111 | 33.64 |  |
| 3 |  | 332 | 29.56 | 133 | 40.3 |  |
| Age years |  | 6.92 | 6.18, 7.61 | 6.93 | 6.10, 7.75 | 0.8923 |
| Body Mass Index Z-score |  | 0.306 | -0.50, 1.31 | 0.27 | -0.57, 1.23 | 0.0414 |

#### Accelerometry outcomes

Physical activity results are reported in Table 3 There was evidence of a meaningful effect of the JU:MP intervention on the primary outcome, average daily habitual MVPA, with an adjusted mean difference of +4.99 minutes/day in the JU:MP group compared to the control group at 24 months follow-up. The effect was stronger on weekdays, with a difference of +5.77 minutes/day (SMD = 0.35) compared to a difference of +4.18 minutes/day on weekends (SMD = 0.15). Secondary accelerometer outcomes showed meaningful differences in average daily ST with a difference of -8.69 minutes/day (SMD = -0.20) between JU:MP and control group at 24 months follow-up and a larger difference observed on weekends (-21.47 minutes/day, SMD = -0.40). The difference in average daily LPA between JU:MP and control groups at 24 months was only different on weekend days +11.75 minutes/day (SMD = 0.29). Additionally, CPM, a measure of total physical activity, improved by 32.72 CPM (SMD = 0.28) overall (JU:MP group compared to control group at 24 month follow-up), with a stronger weekday effect (35.34 CPM, SMD = 0.31).

**Table 3.**
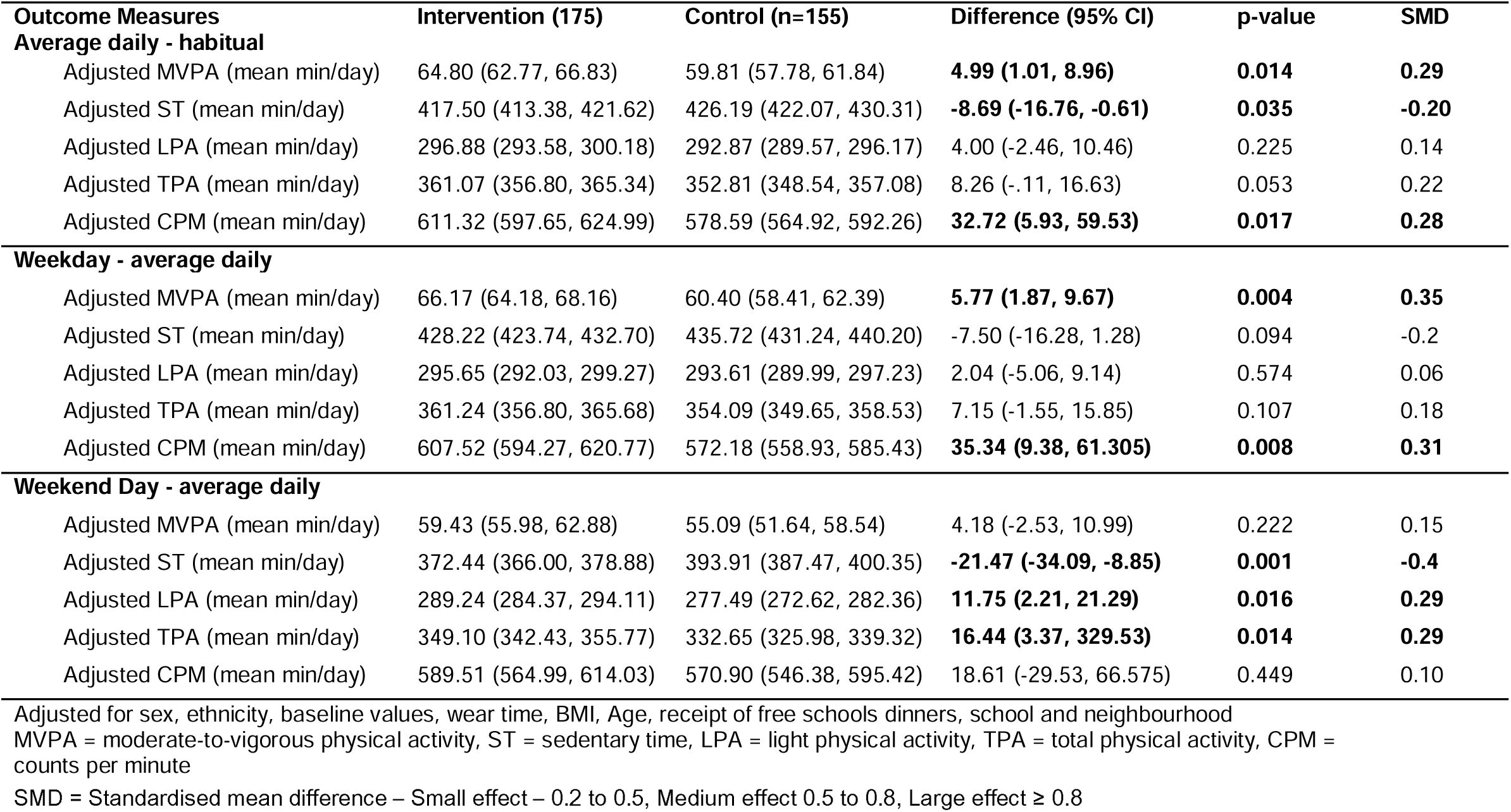
Effectiveness outcomes for average daily habitual behaviours.

| **Outcome Measures** | **Intervention (175)** | **Control (n=155)** | **Difference (95% CI)** | **p-value** | **SMD** |
| --- | --- | --- | --- | --- | --- |
| **Average daily - habitual** |  |  |  |  |  |
| Adjusted MVPA (mean min/day) | 64.80 (62.77, 66.83) | 59.81 (57.78, 61.84) | **4.99 (1.01, 8.96)** | **0.014** | **0.29** |
| Adjusted ST (mean min/day) | 417.50 (413.38, 421.62) | 426.19 (422.07, 430.31) | **-8.69 (-16.76, -0.61)** | **0.035** | **-0.20** |
| Adjusted LPA (mean min/day) | 296.88 (293.58, 300.18) | 292.87 (289.57, 296.17) | 4.00 (-2.46, 10.46) | 0.225 | 0.14 |
| Adjusted TPA (mean min/day) | 361.07 (356.80, 365.34) | 352.81 (348.54, 357.08) | 8.26 (-.11, 16.63) | 0.053 | 0.22 |
| Adjusted CPM (mean min/day) | 611.32 (597.65, 624.99) | 578.59 (564.92, 592.26) | **32.72 (5.93, 59.53)** | **0.017** | **0.28** |
| **Weekday - average daily** |  |  |  |  |  |
| Adjusted MVPA (mean min/day) | 66.17 (64.18, 68.16) | 60.40 (58.41, 62.39) | **5.77 (1.87, 9.67)** | **0.004** | **0.35** |
| Adjusted ST (mean min/day) | 428.22 (423.74, 432.70) | 435.72 (431.24, 440.20) | -7.50 (-16.28, 1.28) | 0.094 | -0.2 |
| Adjusted LPA (mean min/day) | 295.65 (292.03, 299.27) | 293.61 (289.99, 297.23) | 2.04 (-5.06, 9.14) | 0.574 | 0.06 |
| Adjusted TPA (mean min/day) | 361.24 (356.80, 365.68) | 354.09 (349.65, 358.53) | 7.15 (-1.55, 15.85) | 0.107 | 0.18 |
| Adjusted CPM (mean min/day) | 607.52 (594.27, 620.77) | 572.18 (558.93, 585.43) | **35.34 (9.38, 61.305)** | **0.008** | **0.31** |
| **Weekend Day - average daily** |  |  |  |  |  |
| Adjusted MVPA (mean min/day) | 59.43 (55.98, 62.88) | 55.09 (51.64, 58.54) | 4.18 (-2.53, 10.99) | 0.222 | 0.15 |
| Adjusted ST (mean min/day) | 372.44 (366.00, 378.88) | 393.91 (387.47, 400.35) | **-21.47 (-34.09, -8.85)** | **0.001** | **-0.4** |
| Adjusted LPA (mean min/day) | 289.24 (284.37, 294.11) | 277.49 (272.62, 282.36) | **11.75 (2.21, 21.29)** | **0.016** | **0.29** |
| Adjusted TPA (mean min/day) | 349.10 (342.43, 355.77) | 332.65 (325.98, 339.32) | **16.44 (3.37, 329.53)** | **0.014** | **0.29** |
| Adjusted CPM (mean min/day) | 589.51 (564.99, 614.03) | 570.90 (546.38, 595.42) | 18.61 (-29.53, 66.575) | 0.449 | 0.10 |

Adjusted for sex, ethnicity, baseline values, wear time, BMI, Age, receipt of free schools dinners, school and neighbourhood MVPA = moderate-to-vigorous physical activity, ST = sedentary time, LPA = light physical activity, TPA = total physical activity, CPM = counts per minute
SMD = Standardised mean difference – Small effect – 0.2 to 0.5, Medium effect 0.5 to 0.8, Large effect $\geq 0.8$

#### Secondary Outcomes

Secondary outcomes (Table 4) (BMI z-scores, social emotional and behavioral health, and quality-of-life measures) had small, unimportant differences between groups. Exploratory analysis found evidence of meaningful effects from the JU:MP intervention for boys and South Asian children, with MVPA improving for both compared to the control group (+7.34 minutes/days, SMD = 0.36 and +7.20 minutes/day, SMD = 0.52 respectively), there were smaller increases for girls and White British children.

**Table 4.** Effectiveness outcomes for anthropometry, strengths and difficulties and quality of life.

|  |  |  |  |  |  |
| --- | --- | --- | --- | --- | --- |
| **Secondary Outcome Measures** | **Intervention (95% CI)** | **Control (95% CI)** | **Difference (95% CI)** | **p-value** | **SMD** |
| **Anthropometric** |  |  |  |  |  |
| Body Mass Index Z score (n=1,103) | .492 (0.43, 0.55) | .559 (0.49, 0.22) | -.066 (-0.152, .0,211) | 0.138 | 0.09 |
| Waist Circumference inches (n=1098) | 64.08 (63.65, 64.52) | 63.40 (62.69, 64.12) | 0.68 (-0.11, 1.48) | 0.092 | 0.10 |
| **Social, emotional and behaviour difficulties–Teacher Reported SDQ** |  |  |  |  |  |
| Total difficulties score (n=675) | 6.60 (5.08, 7.06) | 6.70 (5.77, 7.63) | -0.634 (-1.99, 0.73) | 0.362 | -0.11 |
| Emotional score (n=676) | 1.60 (1.34, 1.86) | 1.76 (1.50, 2.01) | -0.157 (-0.519, 0.205) | 0.395 | -0.08 |
| Conduct score (n=677) | 0.84 (0.63, 1.04) | 1.03 (0.82, 1.24) | -0.192 (-0.486, 0.100) | 0.199 | -0.13 |
| Hyperactivity score (n=677) | 2.69 (2.27, 3.10) | 2.78 (2.39, 3.18) | -0.098 (-0.67, 0.478) | 0.738 | -0.04 |
| Peer-problem score (n=677) | 0.95 (0.71, 1.19) | 1.23 (1.01, 1.46) | -0.29 (-0.617, 0.042) | 0.087 | -0.19 |
| Prosocial score (n=677) | 8.09 (7.71, 8.48) | 8.34 (7.99, 8.70) | -0.248 (-0.768, 0.272) | 0.350 | -0.13 |
| **Quality of Life – Parent reported** |  |  |  |  |  |
| PEDSQL Score (n=479) | 80.96 (78.40, 83.52) | 79.84 (77.38, 82.31) | 1.12 (-2.44, 4.68) | 0.538 | 0.08 |
| EQ5D Score (n=700) | 0.912 (0.90, 0.93) | 0.915 (0.90, 0.93) | -0.003 (-0.267, 0.020) | 0.783 | -0.03 |

Adjusted for sex, ethnicity, baseline values, wear time, BMI, Age, receipt of free schools dinners, school and neighbourhood
SMD = Standardised mean difference – Small effect – 0.2 to 0.5, Medium effect 0.5 to 0.8, Large effect ≥ 0.8

## Discussion

We report the results of a controlled before and after trial of a whole system approach to improve population-level physical activity that reached over 30,000 children aged five to14 years from a multi-ethnic and economically deprived area. The JU:MP intervention effectively mitigated the age-related decline in MVPA levels, resulting in a relative improvement of five minutes per day compared to the control group after 24 months. The study also found evidence of meaningful differences in average daily TPA, CPM and ST between the JU:MP intervention and control group demonstrating effects of the intervention across multiple movement behaviours. The difference in overall average MVPA was driven by more substantial improvements on weekdays compared to weekends. On weekends, children in the JU:MP group improved their levels of LPA while also reducing ST. While these effects were small (SMD 0.26 to 0.40), at a population level, this demonstrates the substantial public health impact of the JU:MP whole system approach to improve physical activity behaviours within multi-ethnic and socially and economically deprived communities.

Our study found that daily MVPA increased by an average of five minutes, with a noticeable effect size of 0.29, rising to 0.34 on weekdays. Compared to Metcalf et al.’s meta-analysis of children’s physical activity interventions, which reported an effect size (SMD) of 0.16 and approximately four additional minutes of MVPA, our results are promising (51). Specifically, the JU:MP whole system intervention which targets entire child populations (rather than just those with overweight or obesity), shows higher effectiveness than single or multi-component interventions (SMD 0.07 in Metcalf et al. (51)). Our findings also align with another recent meta-analysis (52), indicating a pooled effect size (SMD 0.34) for physical activity interventions among low-income and ethnic minority children. Notably, these interventions tend to be effective over shorter durations (<13 weeks) but struggle to maintain long-term effects, highlighting the JU:MP intervention’s success in fostering behaviour change over two years. Unlike many studies, JU:MP operated as a comprehensive intervention implemented at scale and mitigated common pitfalls such as reduced MVPA levels observed during scaling due to implementation challenges like programme fidelity and adaptability (53). JU:MP’s model included learning cycles, facilitating adjustments to these challenges, potentially overcoming issues encountered in other efficacy or effectiveness studies influenced by external factors like local partnerships, political support, and resource allocation.

Larger improvements in MPVA were observed on weekdays (+5.77 minutes/day), compared to weekends (+4.18 minutes/day). Explanations may be that this is when the children were exposed to physical activity opportunities in schools. Additionally, a large proportion of the children also attended Madrasas on weekdays due to their Muslim faith and the JU:MP intervention increased their physical activity opportunities in these settings too. Our research evidence organisational and cultural change for physical activity, which, in turn, has likely resulted in improved physical activity opportunities for pupils within these settings (54,55). Conversely, the overall differences in TPA and ST were driven by improvements at the weekend (+17.2 minutes and -22.15 minutes difference between groups respectively) suggesting that the workstreams operating at the ‘environment, community and family levels’ where physical activity is less structured were likely responsible for total physical activity and sedentary time improvements. The improvement in weekend TPA was predominantly due to improved LPA (+12.1 minutes difference between groups) which may reflect children swapping ST for light physical activities.

Evidenced by a previous meta-synthesis (56), it is important to understand potential sub-population effects of interventions to ensure equitable benefit and that interventions reach those with greatest health inequality (57). The exploratory analysis found JU:MP had a greater effect on MVPA for boys (+7.3 minutes, SMD 0.36) than girls, and South Asian children (+7.2 minutes, SMD 0.52) than white British children. This indicates that the intervention components activated boys and South Asian children more than girls and White British children. The outcome for South Asian children is positive, given the need to address their lower levels of physical activity compared to White British peers (57). Yet the challenge persists in improving girls’ physical activity. The lack of effect on secondary outcomes suggests that the intervention was not sufficient to change these outcomes within the follow-up period.

The findings demonstrate the value of taking a whole systems approach to improving population levels of physical activity. We have some initial ideas about what the important elements of the intervention might be, based on existing literature. These are: first, the JU:MP intervention had ‘a high use of system approaches’ (according to the analytical framework of Koorts et al. (12)) and activated multiple systems, providing a broad array of physical activities with partners within neighbourhoods and across the Braford city district (22). Second, it was underpinned by theory (complex adaptive systems thinking, the socio-ecological (58) and COM-B behaviour change models (59)) and previous research suggests that theory-based interventions tend to be more effective than those not referencing theory (60). Third, the intervention and its individual elements were co-produced with stakeholders, perhaps enhancing the relevance, engagement, and sustainability of the intervention (61). Fourth, a new district-wide physical activity strategy was co-developed with city-wide partners; we have clear evidence to demonstrate that this helped increase and strengthen relationships between organisations (62). Fifth, national level advocacy and funding (via Sport England) ensured political support for a whole system place-sensitive way of working, with funding to support the development, delivery and evaluation of the intervention (14). Sixth, the iterative learning process embedded within the intervention delivery allowed for continuing adjustments to the intervention and individual elements which may have met practical challenges of implementing interventions in practice and to adapt the intervention to contextual changes (e.g. social, political, economic and research evidence) (63). Alongside the trial, our research group has also conducted a systematic and rigorous process evaluation which will help to unpick whether these intervention elements were important and further explore the mechanism of effects (21,64).

The findings from this study are relevant to public health particularly because there is currently limited evidence for the impact of adopting a systems approach in physical activity interventions (11). This study found that the JU:MP intervention improved MVPA by 5 minutes/day; given the dose-response relationship between physical activity and health outcomes for children and young people, any improvement in physical activity is better than none (65). Moreover, the intervention slowed the rate of age-related decline in physical activity (equivalent to ∼1.6 minutes/day/year) compared to controls (equivalent to ∼3.7 minutes/day/year), even more important given that the JU:MP intervention group had higher levels of deprivation than the control group. This compares favourably to UK and European cohort studies that have shown children’s physical activity levels decline from the age of six to seven through to fourteen to fifteen years at a rate of 2.2 minutes/day/year (66) and ∼2.8 minutes/day/year (1) respectively. As physical activity behaviours in mid-childhood track into adolescence (4), if the effects of JUMP are sustained year-on-year, these could amount to a difference of ∼12 minutes of physical activity per day by the age of fourteen to fifteen. The threshold daily time for MVPA to be health enhancing for children is not clearly defined and is thought to be between 30-60 minutes/day on 3 or more days a week (65). Thus, a possible 12 minute improvement by adolescence at a population level (if behaviours are sustained year on year) could shift a significant number of young people out of the ‘inactive, <30 minutes/day’ zone and into the >30 minutes zone, and achieve important population level public health improvements. Further research would be required to track the young people and assess long-term intervention impact in mid-adolescence to validate these claims.

There is a paucity of detailed descriptions of the operationalisation of whole system public health interventions. JU:MP provides a clear example of an effective whole system approach, with resources detailing the operationalisation of the intervention that local and national governments can learn from (22); as such this research can be viewed as an early step in the development and adoption of effective whole system approaches to physical activity. However, complex whole system interventions cannot simply be transplanted into new localities, decision-makers need to consider the appropriateness of both the intervention components themselves (e.g. the workstreams and their interactions) and how the intervention might interact with the context in which it is being implemented. For researchers, this means evaluations need to be appropriate and multifaceted. Recently released guidance and framework (63,67) suggest that although research should still explore the effectiveness of interventions, a broader range of questions should also be posed, e.g., what are the wider impacts? What is the value relative to the resources to deliver? How does it work, for who and why, taking into account the context in which it is implemented? How does it contribute to system change? The evaluation of JU:MP has taken this multi-faceted approach and has used a mixed methods process evaluation which will explore these questions. Future focuses of research on the JU:MP intervention include, how can the JU:MP intervention be sustained locally, how can the intervention be scaled out to the whole of the Bradford District, what is the impact of this and what is the long-term impact of the intervention upon physical activity and health outcomes? Whether the JU:MP intervention effects can be sustained over the longer-term with lower resources? The trial is undertaking a 36-month follow-up which will further explore this alongside a cost-benefit evaluation of the intervention.

This study has several strengths. To our knowledge it represents the first published effectiveness evaluation of a whole systems approach to increasing children’s physical activity, demonstrating its feasibility despite significant time and resource demands. A pragmatic intention-to-treat design ensured all eligible children within recruited schools were included in the analysis, regardless of their engagement in the JU:MP intervention, thus assessing real-world effectiveness. The use of accelerometry to measure physical activity is a key strength, providing robust and reliable outcome data. The study’s controlled before and after design whilst introducing some bias into the study, allowed for naturalistic evaluation in underserved settings, with matched control neighbourhoods improving comparability. Furthermore, the focus on deprived and ethnically diverse neighbourhoods highlights the study’s relevance for addressing health inequalities, and populations often underrepresented in physical activity interventions.

Limitations include the before and after study design which restricts causal inference due to the absence of randomisation and potential unmeasured confounding. Additionally, individual exposure to the various elements of the JU:MP intervention was not assessed, complicating efforts to isolate specific components driving the observed effects. Furthermore, the geographical focus on underserved areas in Bradford and Yorkshire may limit the generalisability of findings to other populations or regions. Furthermore, only two measurement points, baseline and 24 months, were included, which may have missed short-term impacts or changes. Data completeness was another limitation, with only 22.7% of children meeting accelerometer wear-time criteria at both time points, potentially affecting the interpretation and generalisability of findings. This aligns with previous studies, where accelerometer compliance rates are often lower in socioeconomically deprived populations (68–71). Barriers such as participant burden, device discomfort, and lower parental engagement may have contributed to data loss (43,70,71). Future studies should explore strategies to enhance compliance, including wrist-worn devices, reduced wear-time thresholds, and targeted support. Finally, the study was conducted during the COVID-19 pandemic (recruitment in 2021), which may have influenced physical activity levels and JU:MP intervention delivery.

## Conclusion

The current study presents the findings of the first controlled before and after trial of a whole system, population-level physical activity intervention that reached over 30,000 children aged five to 14 years from a multi-ethnic and economically deprived area. The JU:MP intervention was found to be effective with levels of physical activity in the intervention group higher than the control, suggesting that this systems approach is a promising way of mitigating the age-related decline in moderate to vigorous physical activity in children. Whole systems approaches hold considerable promise for addressing population levels of physical activity, whilst also tackling inequalities and improving public health outcomes at scale.

## Supporting information

Strobe checklist

## Data Availability

The dataset that was generated and analysed during the current study is available from the corresponding author upon reasonable request.

## Acknowledgements

Thank you to Dr Lucy Eddy, Sameera Ali, Shania Boom, for supporting the recruitment and data collection. Thank you, Claire Watson, Cathy Hullin, Dave Turner, Cara Staniforth, Zoe Richardson, Sadaf Bashir, Emma Young, Elliot Lever, Mevagh Harris, Lauren Charlesworth Laurence Ewles, and our wider BiB interns who were involved in the data collection for the study. The study was only possible because of the enthusiasm and commitment of the children, families and schools across Bradford and Yorkshire that participated and the authors sincerely thank these groups for taking part in the study.

## Funding

This study was supported by Sport England’s Local Delivery Pilot - Bradford; weblink:https://www.sportengland.org/campaigns-and-our-work/local-delivery. Sport England is a non-departmental public body under the Department for Digital, Culture, Media and Sport (DCMS). The authors’ involvement was also supported by the National Institute for Health Research (NIHR) Yorkshire and Humber Applied Research Collaboration (ARC) the Bristol Biomedical Research Centre (BRC) and the UK Prevention Research Partnership (MR/S037527/1), an initiative funded by UK Research and Innovation Councils, the Department of Health and Social Care (England) and the UK devolved administrations, and leading health research charities. Weblink: https://mrc.ukri.org/research/initiatives/prevention-research/ukprp/. The views expressed in this publication are those of the author(s) and not necessarily those of Sport England, the DCMS, or the NIHR ARC, BRC or UKPRP.

## Contributions

SEB and DB are co-lead authors, contributing equally with varied input at each study stage. The study was conceived by SEB, DB, and ADS. DB led the protocol registration, ethics approval, data processing, and cleaning. Recruitment and data collection were led by SEB, DB and ZH. DB and GS conducted the statistical analysis. SEB, NPD, DB, and ADS drafted the manuscript, with SEB and DB revising it following input from all authors. All authors approved the final manuscript.

## Conflicts of interest

The JU:MP whole system approach was delivered by a consortium of partners on behalf of Active Bradford funded by Sport England. During the study period, Active Bradford was a not-for-profit company limited by guarantee. In 2024 Active Bradford became a charity and from 2025 will host the JU:MP approach. SEB and JW sit on the Directors and Members Boards of Active Bradford respectively. JW was not directly involved in the development or delivery of the intervention. SEB, JH, AS and ADS were involved in the evidence-based development of JU:MP but not the operational delivery of the intervention. JB was the implementation Director for the JU:MP whole system approach and was directly responsible for the delivery of JU:MP. To ensure independence, JB was not involved in the collection or analysis of the data. DB, ZH, GS, and RRCM declare no conflicts of interest.

## Notes

### Clinical Trial

ISRCTN14332797

### Funding Statement

This study was funded by Sport Englands Local Delivery Pilot. Sport England is a non-departmental public body under the Department for Digital Culture Media and Sport (DCMS). The authors involvement was also supported by the National Institute for Health Research (NIHR) Yorkshire and Humber Applied Research Collaboration (ARC) the Bristol Biomedical Research Centre (BRC) and the UK Prevention Research Partnership (MRS0375271) an initiative funded by UK Research and Innovation Councils the Department of Health and Social Care (England) and the UK devolved administrations and leading health research charities.The views expressed in this publication are those of the authors and not necessarily those of Sport England, the DCMS or the NIHR ARC BRC or UKPRP.

### Author Declarations

The study and all processes were given ethical approval by the University of Bradford Research Ethics Committee (Humanities, Social, and Health Sciences Research Ethics Panel; reference: (E891, July 2021).

