## Supplementary Table 1 for "The Effectiveness of a Whole System Approach to Improve Physical Activity of Children Aged 5 to 11 Years Living in Multi-ethnic and Socio-economically Deprived Communities: A Controlled Before and After Trial"

Supplementary file 1: Characteristics and descriptives of recruited schools and neighbourhoods

|  | No. Schools | No. pupils in Years 1 to 3 | Median % of pupils known to be eligible for free school meals | Median % of pupils classified as White British | Median % of pupils classified as South Asian | Median School IMD* decile |
| --- | --- | --- | --- | --- | --- | --- |
| Neighbourhood 1 |  |  |  |  |  |  |
| Intervention | 6 | 789 | 25.9 | 73.15 | 10.45 | 2.5 |
| Control | 7 | 718 | 34.9 | 70.9 | 9.15 | 2 |
| Total | 13 | 1507 | 28.9 | 72.1 | 9.65 | 2 |
| **Difference** | **-1** | **71** | **-9** | **2.25** | **1.3** | **0.5** |
| Neighbourhood 2 |  |  |  |  |  |  |
| Intervention | 5 | 840 | 30.6 | 0.8 | 86.2 | 1 |
| Control | 7 | 798 | 22.1 | 3.2 | 85.5 | 2 |
| Total | 12 | 1638 | 24 | 1.55 | 85.85 | 1.5 |
| **Difference** | **-2** | **42** | **8.5** | **-2.4** | **0.7** | **-1** |
| Neighbourhood 3 |  |  |  |  |  |  |
| Intervention | 6 | 613 | 42.1 | 68.75 | 2.3 | 1.5 |
| Control | 6 | 650 | 37 | 81.9 | 2.45 | 1.5 |
| Total | 12 | 1263 | 42.1 | 80.5 | 2.3 | 1.5 |
| **Difference** | **0** | **-37** | **5.1** | **-13.15** | **-0.15** | **0** |
| Total |  |  |  |  |  |  |
| Intervention | 17 | 2242 | 30.7 | 57.8 | 17.5 | 1 |
| Control | 20 | 2166 | 26.6 | 61.35 | 17.9 | 2 |
| Total | 37 | 4408 | 30 | 57.8 | 17.5 | 1 |
| **Difference** | **-3** | **76** | **4.1** | **-3.55** | **-0.4** | **-1** |
| Source of data was Department of Education Census Academic year 2021/22 - Schools, pupils and their characteristics  <https://explore-education-statistics.service.gov.uk/find-statistics/school-pupils-and-their-characteristics/2021-22>  * IMD = Index of Multiple Deprivation, based upon school postcode | | | | | | |
